# Evaluating sustained reach and effectiveness of collaborative care models: A Cross-sectional study of the New York State Collaborative Care Medicaid Program

**DOI:** 10.1101/2025.01.11.25320387

**Authors:** Kain Kim, Baoyi Feng, Mengxiao Luan, Jungang Zou, Amy Jones, Danielle Gadbois, Joseph E. Schwartz, Qixuan Chen, Nathalie Moise

## Abstract

**Background:** Little data exists on collaborative care (CC) sustainability.

**Objective:** Describe and determine predictors of long-term CC reach and effectiveness Design: Cross-sectional observational study of the NY State CC Medicaid Program (CCMP), involving technical assistance (TA), quality monitoring, and fee-for-quality Medicaid reimbursement codes for implementing CC. We included clinics participating in CCMP from 2012-2019 with ≥0.5 full time equivalent (FTE) care manager and available 2021 and/or 2021 data.

**Main Measures:** Clinic (size, type, region, enrollment year); and CC program (care manager FTE, caseload/care manager FTE [target 100-150], screening [proportion of clinic screened for depression], engagement [proportion of CC patients contacted/engaged monthly], and psychiatrist consultations in unremitted patients) characteristics. Outcomes were reach (proportion of screen-detected depressed patients enrolled in CC) and effectiveness (proportion of CC enrolled patients achieving remission or 50% reduction in depressive symptoms [Target 50-60%]). We used multilevel negative binomial regression models, adjusting for clustering by healthcare system and county.

**Results:** Of eligible 160 clinics, 71.2% were Federally Qualified Health Centers (FQHCs); the median caseload/care manager FTE was 55.1, reach 13.0% and effectiveness 42.0%. In multivariable analyses, key CC factors associated with reach included engagement (adjusted Rate Ratio [aRR]=3.99 [1.82, 8.76]), care manager FTE (aRR=1.06 [1.02, 1.10]), and caseload/care manager FTE (aRR=1.23 [1.17, 1.29]); smaller clinic size (aRR=0.60 [0.53, 0.69]), earlier adoption (aRR=0.40 [0.23,0.69] in 2017-2019 vs. 2012-2014), and academic/private clinics (vs. FHQC) (aRR=0.66 [0.45, 0.96]) were also predictive. Caseload/care manager FTE (aRR=1.04 [1.01, 1.07]), psychiatry consultations (aRR=1.55 [1.19, 2.00]), and FQHCs (aRR=1.19 [1.02, 1.40]) were associated with greater CC effectiveness.

**Conclusion:** Despite ongoing fiscal and TA, CC clinics particularly struggle to achieve long-term reach. While majority FQHCs limit generalizability, we provide several targets for selecting ideal settings for CC, optimizing the pace of sustainability and considering de-implementation efforts when futile.

**Primary Funding Source:** Agency for Healthcare Research

## Background

The collaborative care model for depression (CC) is an integrated behavioral health model shown to improve access to mental health resources, depression symptoms, and quality of life compared to usual primary care, and is one of the few evidence-based interventions in general shown to reduce health disparities.^1–6^ Over the last decade, experts have sought to target the wide range of implementation barriers,^1,7–10^ with fiscal, training, and implementation support strategies showing wide ranging results.^11–13^ CC is now widely reimbursed and it is now clear that that achieving real-world effectiveness similar to clinical trials is contingent on the level of external implementation support.^1^

Despite remarkable progress in CC implementation, there has been little focus on sustainability,^14–16^ defined as “the continued use of program components and activities for the *continued achievement of desirable program and population outcomes.*”^17^ The key determinants of sustained reach (i.e., percentage of eligible patients enrolled in CC) and effectiveness (i.e., percentage of enrolled patients with remission and/or >=50% reduction in depressive symptoms) remain unknown. In 2015, the New York Office of Mental Health (OMH) launched the Collaborative Care Medicaid Program (CCMP) (implementation strategy), which supported CC (the evidence-based intervention) implementation in primary care clinics across the state with training, centralized technical assistance (TA), ongoing quality monitoring and Medicaid reimbursement codes. The program resulted in broad CC adoption and fidelity, reaching the majority of Federally Qualified Health Centers (FQHCs) in NYS.^18^ To optimize the pace of national CC sustainability efforts and advance the field of implementation science, we aimed to describe and investigate key predictors of sustainability indices in clinics participating in the NYS CCMP.

## Methods

### Setting and Participants

During 2012-2014, the NYS OMH implemented the NYS CC Initiative, a demonstration program funded by the NYS Department of Health.^19^ Having demonstrated improved outcomes in 32 early adopter clinics, OMH launched CCMP in 2015, which became the first Medicaid program in the US to reimburse CC services.^19^ Over 300 clinics currently participate in CCMP, which provides clinics a monthly case rate payment for each Medicaid patient enrolled in CC. A unique feature of CCMP is its adoption of the Quality Supplemental Payment model, which incentivizes clinics to receive additional funding if benchmarks in pre-specified quality outcomes are met.

To participate, primary care clinics were required to submit a billing application to OMH confirming the necessary infrastructure for CC implementation, such as a state-approved registry. Once approved, clinics were required to submit quarterly process and outcomes data used to further improve training and TA. Participating clinics received free training, TA, registry upkeep, and quality improvement activities.^19^ In our study, we included all primary care sites that (1) reported ≥1 quarter’s worth of data in 2021 and/or 2022, (2) participated in NYS CCMP in 2012-2019 to mitigate unique COVID19 effects and isolate sustained programmatic effects, and (3) had ≥0.5 FTE of a care manager.

### Measures and Outcomes

To frame presentation of our data, we employed the Reach, Effectiveness, Adoption, Implementation, and Maintenance (RE-AIM) model.^20^ Our primary outcomes corresponded with the long-term (“maintenance”) constructs of “reach” and “effectiveness” within RE-AIM. Our data source consisted of performance metrics reported on a monthly basis and collected from clinics by OMH quarterly. Our Institutional Review Board at Columbia University Irving Medical Center approved analyses of this quality improvement data.

We calculated reach as the total sum of patients newly enrolled in CC in a given year divided by the number of unique patients who screened positive for depression (defined as a PHQ-9 score of 10 or higher) in that year; only the months with complete data were used. There were rare cases in which the enrollment number exceeded the screen positive number (e.g., the care manager may have enrolled a patient screened in a prior time period or not screened at all), in which case we modified the denominator to equal the numerator, thereby capping reach at 100%. Clinical effectiveness was reported by clinics and defined by OMH (with an optimal goal of 50-60%) as the proportion of patients enrolled in CC for 70 days or greater who demonstrated clinically significant improvement defined as either (1) a PHQ-9 ≤10 or (2) a 50% reduction from their screening/baseline PHQ-9. We calculated effectiveness similar to reach for a given year.

Candidate predictors included various OMH defined metrics related to clinic and CC program characteristics: start year (the year OMH received a complete CCMP application from a clinic, including 2012-2014 (early adopters in the CC Initiative) 2015-2016 and 2017-2019, geographical region (a binary variable of NYC vs. elsewhere in NYS), FQHC status, clinic size (the average monthly number of unique screen eligible patients seen in a clinic), the average monthly care manager FTE, and the average caseload per care manager FTE.^21^ Candidate predictors operationalized similar to reach above (summed numerator over summed denominator in given year) included depression screening (the proportion of all unique adult patients seen during the reporting period who received their annual PHQ-2 or PHQ-9 screening; this did not include patients being monitored for depression as part of CC), engagement (the proportion of CC enrolled patients who had at least one clinical contact with a care manager in a given month), and psychiatry consultation (defined in 2021 as the proportion of those patients enrolled for ≥70 days without improvement who either [1] had their case reviewed by the consulting psychiatrist, who then made treatment recommendations to the PCP or depression care manager, or [2] had a documented change in their treatment plan for that month; in 2022, the second criterion was removed by the OMH due to difficulty capturing data in registries). OMH definitions and targets are included in the Supplementary eText 1.

### Data Analysis

The distributions of outcomes and predictors were examined separately for 2021 and 2022. We present medians and interquartile ranges (IQR, first and third quartiles) for numeric variables and counts with frequencies for categorical variables. We present boxplots to visually compare the 2021 and 2022 depression screening, CC engagement, effectiveness, psychiatrist consultation, and reach.

To investigate the associations between reach/effectiveness and clinic-level characteristics, we fit Bayesian multilevel negative binomial regression models. The models used random intercepts to account for correlations among clinics within the same county, correlations among clinics from the same health systems, and correlations of multiple measures from the same clinics. We started with univariable models examining each predictor separately, followed by multivariable models that included all predictors. Experts posit that reach and effectiveness may be negatively associated.^22^ As such, in the models predicting reach, effectiveness was included as a covariate, and in the models predicting effectiveness, reach was included as a covariate. To reduce the skewness of the covariates, clinic size was log-transformation (natural log) and caseload per care manager FTE was square root transformed. We implemented the Bayesian multilevel negative binomial models using the “*stan_glmer.nb*” function in the “*rstanarm*” package (version 2.32.1) in R. We used the default prior settings in the function and specified 2 chains, each with 4,000 iterations for the burn-in phase and 4,000 for the posterior sampling phase. We monitored the convergence of MCMC chains to obtain a R-hat statistic close to 1. The regression coefficients were obtained using posterior medians and 95% credible intervals (CIs), defined using the 2.5^th^ and 97.5^th^ percentile of the posterior draws. The regression coefficient estimates were then exponentiated to obtain rate ratios (RRs). The 95% CI that does not include 1 was considered statistically significant.

## Results

In total, 160 CCMP clinics met eligibility criteria (109 reported data in both years; 42 in 2021 only, 9 in 2022 only). Table 1 shows similar descriptive statistics in years 2021 and 2022. In 2022, CCMP clinics served a median of 474 [IQR: 159 - 938] patients per month; 56.8% were in NYC; 71.2% FQHCs. Median reach (i.e., the % of patients screened positive who enrolled in CC) was 13% (IQR 5% - 22%), clinical effectiveness 42% (IQR: 26% - 53%), caseload/FTE 55 (30 - 89), depression screening 81% (67% - 94%), engagement (i.e., proportion of enrolled patients with clinical encounters) 58% (42% - 70%) and psychiatry consultation 38% (11% - 71%). Figure 1 displays the distributions of depression screening, reach, engagement, effectiveness, and psychiatric consultation in 2021 and 2022 using boxplots. Figures 2 and 3 show the average reach and effectiveness for each county.

**Figure 1.**
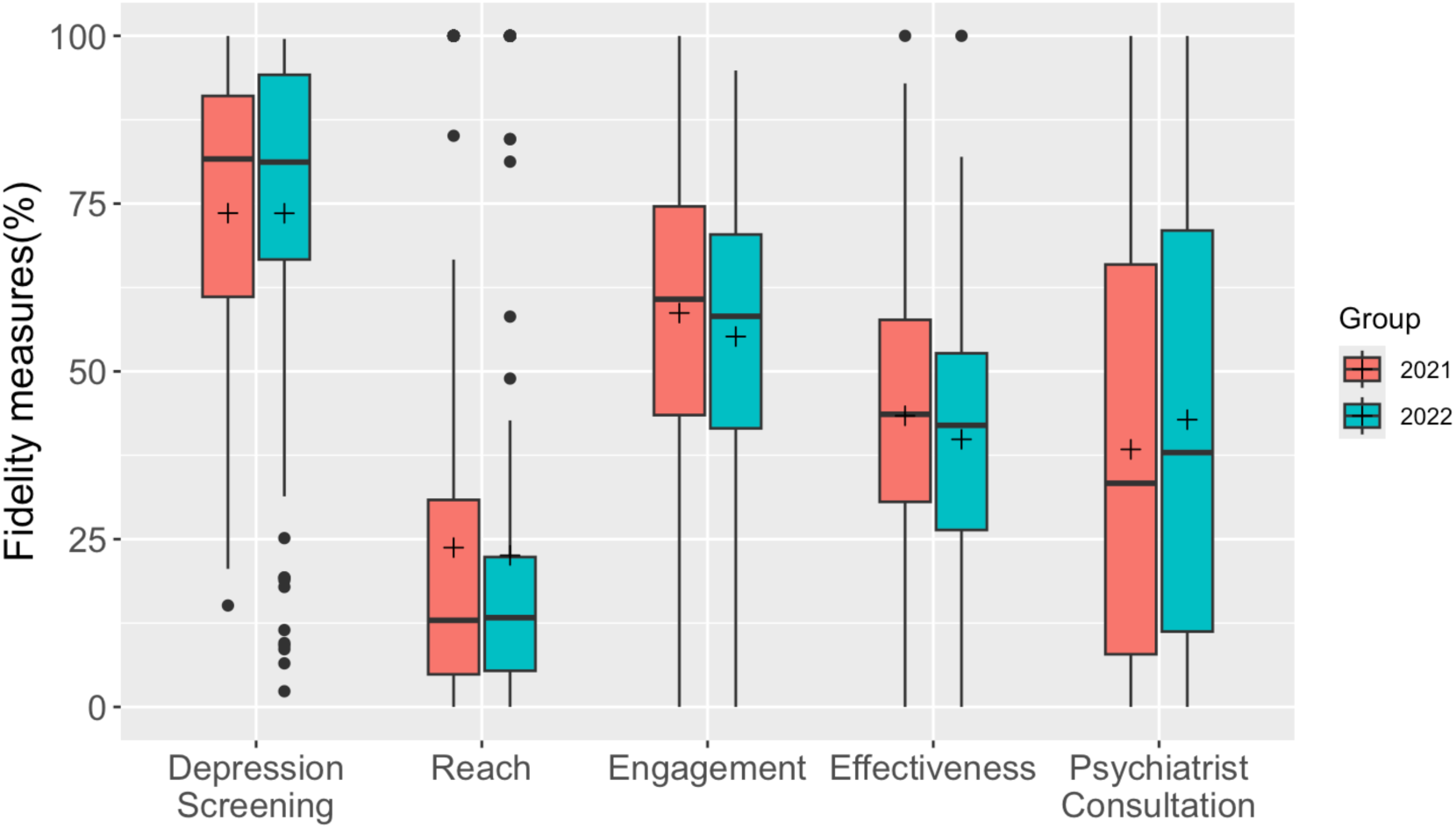
Comparing fidelity measures between 2021 and 2022 using boxplots: + sign representing sample mean

**Table 1.** Descriptive statistics of outcomes and covariates for the clinics with data in 2021 or 2022: count (percentage) for categorical variables and median (Q1, Q3) for numerical variables.

|  | 2021 (n = 151) | 2022 (n = 118) |
| --- | --- | --- |
| Starting year |  |  |
| 2012-2014 | 18 (11.9%) | 17 (14.4%) |
| 2015-2016 | 24 (15.9%) | 19 (16.1%) |
| 2017-2019 | 109 (72.2%) | 82 (69.5%) |
| Region |  |  |
| New York City | 78 (51.7%) | 67 (56.8%) |
| Other regions | 73 (48.3%) | 51 (43.2%) |
| Federally Qualified Health Centers (FQHC) |  |  |
| FQHC | 111 (73.5%) | 84 (71.2%) |
| Not FQHC | 40 (26.5%) | 34 (28.8%) |
| Clinical size | 444 (124, 963) | 474 (159, 938) |
| Caseload per FTE | 50 (25,74) | 55 (30,89) |
| Care manager FTE | 1 (0.68,1.24) | 1 (0.61,1.28) |
| Reach | 13% (5%, 31%) | 13% (5%, 22%) |
| Effectiveness<sup>a</sup> | 44% (31%, 58%) | 42% (26%, 53%) |
| Engagement | 61% (43%, 75%) | 58% (42%, 70%) |
| Psychiatry consultation<sup>b</sup> | 33% (8%, 66%) | 38% (11%, 71%) |
| Depression screening | 82% (61%, 91%) | 81% (67%, 94%) |

<sup>a</sup> 4 clinics did not have effectiveness data in 2021
<sup>b</sup> 5 clinics did not have psychiatry consultation data in 2021 and 1 clinic in 2022

Table 2 shows the results of the multivariable models using Bayesian multilevel negative binomial regression for reach and effectiveness. After adjusting for other covariates, there was no difference in reach between 2021 and 2022 or between clinics in NYC versus other regions in NYS. Reach was associated with earlier adoption (2017-2019: aRR=0.40 [95% CI: 0.23, 0.69], 2012-2014: reference), smaller clinic size (aRR=0.60 [95% CI: 0.53, 0.69]), larger caseload per care manager FTE (aRR=1.23 [95% CI: 1.17, 1.29]), higher care manager FTE (aRR=1.06 [95% CI: 1.02, 1.10]), better engagement (i.e., proportion of enrolled patients with care manager clinical contact) efforts (aRR=3.99 [95% CI: 1.82, 8.76]), academic/private clinic status (aRR=0.66 [95% CI: 0.45,0.96]) and lower effectiveness (aRR=0.47 [95% CI: 0.25, 0.91]), but not psychiatrist consultation or depression screening metrics. Effectiveness was associated with higher caseload per care manager FTE (aRR=1.04 [95% CI: 1.01, 1.07]), greater psychiatrist consultations for unremitting patients (aRR=1.55 [95% CI: 1.19, 2.00]), FQHC clinic status (aRR=1.19 [95% CI: 1.02,1.40]), and lower reach (aRR: 0.56 [0.41, 0.77]). The results of the univariable models are presented in the supplementary eTable 1.

**Table 2.** Bayesian multivariable multilevel negative binomial regression models examining factors associated with reach and effectiveness among CCMP clinics starting before 2020. The rate ratio (RR) and 95% credible interval (CI) are presented.

|  | Reach<br>RR (95% CI) | Effectiveness<br>RR (95% CI) |
| --- | --- | --- |
| Year of data: 2022 vs 2021 | 1.01 (0.88,1.16) | 0.91 (0.82,1.00) |
| Region: NYC vs other | 1.23 (0.44,3.39) | 1.08 (0.81,1.48) |
| Starting year (Ref = 2014) |  |  |
| 2015-2016 | 0.56 (0.31,1.01) | 0.92 (0.71,1.18) |
| 2017-2019 | 0.40 (0.23,0.69) | 0.79 (0.62,0.99) |
| Log-transformed clinic size | 0.60 (0.53,0.69) | 0.97 (0.91,1.04) |
| Sqrt transformed caseload per FTE | 1.23 (1.17,1.29) | 1.04 (1.01,1.07) |
| Care manager FTE | 1.06 (1.02,1.10) | 1.01 (0.99,1.03) |
| Engagement | 3.99 (1.82,8.76) | 1.29 (0.86,1.92) |
| Psychiatry consultation | 0.91 (0.57,1.44) | 1.55 (1.19,2.00) |
| Depression screening | 1.28 (0.65,2.58) | 0.79 (0.56,1.13) |
| FQHC | 0.66 (0.45,0.96) | 1.19 (1.02,1.40) |
| Effectiveness | 0.47 (0.25,0.91) | - |
| Reach | - | 0.56 (0.41,0.77) |

## Discussion

The purpose of this study was to describe and understand determinants of long-term effectiveness and reach of the NYS CCMP, one of the largest statewide programs of its kind. Informed by the RE-AIM framework, our study demonstrated that despite ongoing fiscal and technical assistance strategies, clinics struggled with sustained reach, and to a lesser extent, effectiveness. Our findings also suggest a possible tradeoff between reach and clinical effectiveness, both of which were associated with caseload/care manager FTE, a potential target for policymakers seeking to sustain CC programs.

To date, researchers have largely focused on effectiveness and less so reach of CC programs. Experts posit that achieving population wide impact may require maximizing reach even while marginally sacrificing effectiveness.^22^ Here, we found that significant predictors of long-term reach included better patient engagement efforts and care manager FTE (unadjusted for caseload) both of which should be targeted to optimize the pace of sustainability efforts. A multiple case study of the collaborative chronic care model in outpatient mental health clinics highlighted the need for increased prioritization of fundamental operational needs (e.g. mental health staffing or clinical space).^23^ These resources remain integral in the sustainability phase when psychosocially complex patients and their providers become increasingly difficult to reach, particularly in FQHCs. Clinics seeking to sustain CC may address resource barriers by task-shifting duties to lower-cost administrative roles to expand care manager capacities.^14^ Centralized triage and referral services may also support CC implementation efforts and maximize reach while also meeting the needs of the diverse array of patients, many of whom may require more intensive mental health services.^24^ Our study results also provide valuable insights for policymakers, highlighting the need to target efforts toward specific settings, such as larger clinics, later adopters, and Federally Qualified Health Centers (FQHCs) that may have fewer resources per capita and which face greater challenges with achieving reach.

Additionally, our study adds to our understanding of the drivers of sustained clinical effectiveness, which has been shown to be particularly difficult to maintain after removing initial implementation support.^17,25–28^ Unutzer found that CC clinics receiving enhanced (vs. basic) implementation support are more likely to achieve the effects of clinical trials (i.e., optimally ≥50-60%).^1^ We advance this literature by demonstrating that despite *sustained implementation support*, clinics see long-term effectiveness rates of 42% which aligns with real world effectiveness rates in CC settings with varying degrees of support.^1^ Here, we found that a higher percentage of psychiatric consultations for unremitted patients (done in just 30-40% of patients) was associated with higher effectiveness, supporting the integral role of psychiatrists, even with case-based consultation, in achieving optimal outcomes.^29^ Psychiatry involvement, which may see a voltage drop in the sustainability phase, should be further incentivized and revived in addition to promoting engagement with external implementation support.

Our results also support recent research suggesting a reciprocal relationship between clinical effectiveness and reach.^22^ This was perhaps due to a decrease in quality that may occur when too many patients are reached and, vice versa, the high degree of effectiveness that can be achieved when care managers focus on only a small group of patients. Our study further highlights the need to explicitly distinguish reach and effectiveness as unique targets in achieving sustainability. For example, capacity building, care manager training/accountability and primary care referral support may improve reach, while maximizing use of e-consults may be an avenue for increasing psychiatrist involvement. Interestingly, greater caseload/care manager FTE, which may serve as a proxy for high-quality care managers, was positively associated with both reach and effectiveness. Notably, median caseload per FTE in our study was approximately half the recommended acuity-adjusted panel size of 100-150 patients/FTE for moderately complex patients.^30^ Targeting caseloads, perhaps through fee-for-quality reimbursement codes that have thus far been focused on incentivizing effectiveness, may be one approach.

Finally, it has now been 10 years since the 2012-2014 OMH/DOH demonstration project found an average effectiveness of 45% and reach (defined as proportion of clinically diagnosed patients, which made up the majority of those screened, enrolled in CC) of 43%.^19,31^ Our study suggests that reach may wane over time, perhaps due to the myriad of unique barriers to CC sustainability.^15,32^ ^53,54^ ^14,33^ Future research is warranted to examine how best to promote reach and effectiveness long-term. It may be that CC is not be worth the effort in every setting, and identifying settings best suited for CC, perhaps via predictive modeling, while de-implementing ineffective models in others is an area of future research.

There were several limitations to this study’s design that restricted our ability to generalize our findings to other states and clinics. First, the majority of our clinics were FQHCs that served Medicaid populations, potentially limiting generalizability. As the available data were collected primarily for administrative purposes and reviewed retrospectively, several variables (e.g., reach) were based on clinic self-report. However, we did mostly draw from OMH’s prespecified definitions of key metrics. Not every clinic participating in CCMP reported data to OMH, and among those reporting data, there was considerable missing data (e.g., in a given month no patients were enrolled or eligible for clinical improvement assessment or EMR/registry issues emerged). As we calculated reach based on the number of enrolled patients divided by the number who screened positive, our data may have been skewed when patients were screened one month and enrolled in another and does not account for screen positive patients who were not eligible for CC (e.g., severe psychiatric comorbidities though care managers often assist in care coordination for such patients). Finally, we could not examine patient-level characteristics such as gender, age and symptom severity due to the lack of patient-level data,^1^ though this Medicaid program has a large proportion of historically marginalized patients. Despite limitations, the outcomes-driven approach of the NYS CCMP provides an opportunity to address the challenges of implementing and sustaining CC in diverse patient population.

## Conclusion

Overall, our study adds to our understanding of the drivers of sustained reach and effectiveness in the context of ongoing implementation support and provides several targets for future interventions. Our study is also a call to action for increased attention to reach, particularly given its importance in achieving equitable access to CC. Large state-sponsored initiatives like NYS’ CCMP will continue to be integral to tackling the challenging work involved in implementing and sustaining CC in diverse patient populations and settings while also advancing our understanding of what it takes to sustain evidence-based programs, which is particularly salient to the emerging field of implementation science.

## Data Availability

The senior author (NM) had full access to the data and takes responsibility for the integrity of the data and the accuracy of the data analysis. The datasets used and/or analyzed in this study are available in Open Science, https://www.cos.io upon request.

## Acknowledgments

The authors would like to thank Drs. Ian Kronish and Jennifer M. Barbecho for their critical revisions to this work.

## Authors’ contributions

KK wrote the first draft of the manuscript. BF, ML, JZ, QC, and JES managed, analyzed and interpreted the data. JZ and QC drafted the data analysis and results sections. NM conceptualized the project, obtained funding and was a key contributor in writing the manuscript. AJ and DG collected and cleaned the data. All authors contributed to critical revision of the manuscript. All authors read and approved the final manuscript.

## Prior presentations

This manuscript has not been previously presented or published elsewhere.

## Ethics approval and consent to participate

This study was approved by the Columbia University Institutional Review Board (study ID: AAAQ1705).

## Consent for publication

Not applicable. Only aggregate clinic-level data (no patient level data) were used in this study.

