## Supplemental Materials for "Evaluating sustained reach and effectiveness of collaborative care models: A Cross-sectional study of the New York State Collaborative Care Medicaid Program"

**# of eText: 1**

**# of eTables: 1**

**Supplementary eText 1. New York State CCMP metrics definitions.**

1. Total Enrollment – The total number of patients enrolled in Collaborative Care treatment during this month (Optimal caseload >75 patients/ 1.0 FTE BH CM)

2. Medicaid Enrollment – The total number of Medicaid patients enrolled in Collaborative Care treatment during this month

3. Newly enrolled – Among enrolled patients, the number of patients who were diagnosed with Depression or Anxiety and enrolled in treatment by the BHCM this month (Optimal rate: 10% of total caseload)

4. Average Duration of Treatment – Average number of weeks between initial assessment to date of discharge from Collaborative Care

5. Monthly Contact – Number (#) and proportion (%) of patients receiving active treatment in Collaborative Care defined by those patients who have had at a clinical contact this month.

Numerator: Patients that have had at least one clinical contact this month

Denominator: Total number of patients enrolled during this month
Note: A “clinical contact” is defined as a contact in which monitoring may occur and treatment is delivered with corroborating documentation in the patient chart. This includes individual or group psychotherapy visits and telephonic engagement as long as treatment is delivered. (2022 Target Rate: ≥80%)

6. Improvement Rate – Number (#) and proportion (%) of patients enrolled in treatment for 70 days or greater who demonstrated clinically significant improvement either by: a. A 50% reduction from baseline PHQ-9/ GAD-7 or b. A drop from baseline PHQ-9/ GAD-7 to less than 10 (2022 Target Rate ≥50%; Optimal Performance Target: ≥60%)

Numerator: Patients that have met Improvement criteria

Denominator: All patients enrolled in Collaborative Care for 70 days or more

7. Remission Rate – Number (#) and proportion (%) of patients enrolled in treatment for any length of time who have achieved remission criteria (PHQ-9/ GAD-7 below 5) during this month

Numerator: Patients whose most recent PHQ-9/GAD-7 is below 5

Denominator: Total number patients enrolled during this month

8. Psychiatric Consultation: Among those enrolled in treatment for 70 days or more who did not improve, number (#) and proportion (%) who whose case was reviewed by the Consulting Psychiatrist with treatment recommendations provided to the Primary Care Provider or Depression Care Manager in the past 60 days. (2022 Target Rate ≥80%)

Numerator: Patients who have had their case reviewed by the Consulting Psychiatrist in the past 60 days

Denominator: Patients that have been enrolled for 70 days or more who have not met clinical improvement criteria this month

The following metrics are reported for your entire patient population, not just Medicaid patients or those actively enrolled in Collaborative Care. These are not in CMTS.

9. Depression Screening Rate – Number (#) and proportion (%) of all unique patients seen during the reporting period who received their annual PHQ-2 or PHQ-9 screening. (*all payers) (2022 Target Rate 85%)

Numerator: Patients that received a PHQ-2 or 9 during this visit, or have been screened in the last year

Denominator: All patients seen in the practice for any reason that month

10. Depression Screening Yield – Number (#) and proportion (%) of all unique adult patients who scored a 10 or greater on their initial PHQ-9 during the reporting period (*all payers)

Numerator: Patients that scored a 10 or higher on their initial PHQ-9

Denominator: All patients screened with a PHQ during that month

11. Behavioral Health Care Manager Staffing – The total FTE devoted to Collaborative Care treatment during this month

Supplementary eTable 1. Bayesian univariable multilevel negative binomial regression models examining factors associated with reach and effectiveness among CCMP clinics starting before 2020. The rate ratio (RR) and 95% credible interval (CI) are presented.

|  | Reach | Effectiveness |
| --- | --- | --- |
|  | RR (95% CI) | RR (95% CI) |
| Year of data: 2022 vs 2021 | 1.04 (0.90,1.21) | 0.92 (0.84,1.01) |
| Region: NYC vs other | 0.73 (0.22,2.43) | 1.28 (0.90,1.84) |
| Starting year (Ref = 2014) |  |  |
| 2015-2016 | 0.40 (0.19,0.83) | 0.89 (0.69,1.15) |
| 2017-2019 | 0.64 (0.35,1.19) | 0.68 (0.54,0.84) |
| Log-transformed clinic size | 0.68 (0.60,0.78) | 1.07 (1.01,1.13) |
| Sqrt transformed caseload per FTE | 1.19 (1.13,1.25) | 1.02 (0.99,1.05) |
| Care manager FTE | 1.01 (0.96,1.05) | 1.01 (0.99,1.03) |
| Engagement | 1.58 (0.74,3.35) | 1.79 (1.27,2.52) |
| Psychiatry consultation | 1.59 (1.00,2.54) | 1.73 (1.40,2.15) |
| Depression screening | 0.45 (0.22,0.90) | 1.04 (0.75,1.43) |
| FQHC | 0.43 (0.28,0.68) | 1.19 (1.00,1.42) |
| Effectiveness  Reach | 0.85 (0.43,1.62) - | - 0.62 (0.46,0.82) |
